# Machine learning fairness analysis on clinical data of The EMory BrEast imaging Dataset (EMBED)

**DOI:** 10.1101/2023.07.23.23293043

**Authors:** Vikas Ramachandra, Mohit Sethi

## Abstract

This paper explores the use of machine learning (ML) for predictive modeling on clinical data from the EMory BrEast imaging Dataset (EMBED) [1] with a focus on ML model fairness analysis. The aim of this study is to develop and evaluate fair machine learning models that can accurately predict breast cancer risk. We trained and tested various machine-learning models. Our findings show that machine learning can be effective for predicting breast cancer risk or diagnosing breast cancer, and that fairness considerations are crucial in the development of such models. Overall, our study highlights the potential of machine learning for clinical applications while emphasizing the need for ethical and fair practices in this field.

## Introduction

Breast cancer is one of the most common forms of cancer in women worldwide. Early detection and accurate diagnosis play a crucial role in the successful treatment of breast cancer. Advances in medical imaging technologies have led to the creation of large datasets of clinical images and associated data. Machine learning (ML) models can be used to analyze such datasets and develop predictive models for the early detection and diagnosis of breast cancer.

The Emory Breast Imaging Dataset (EMBED) is a large and diverse clinical dataset that contains thousands of mammograms, clinical images, and associated data, including patient demographics, mammography findings, pathology results, and treatment outcomes. EMBED provides a valuable resource for researchers and clinicians to develop and test ML-based predictive models for breast cancer diagnosis and treatment.

However, the use of ML models in clinical settings raises concerns about fairness and bias. Models that are trained on biased or unrepresentative data can produce unfair predictions that disproportionately harm certain groups of patients. Therefore, it is important to evaluate the fairness and bias of ML models before deploying them in clinical settings.

In this paper, we describe the development and evaluation of ML-based predictive models on the EMBED dataset for breast cancer diagnosis and treatment. We also perform a comprehensive fairness analysis to ensure that the models are fair and unbiased across different demographic groups. We hope our findings will contribute to the development of more accurate and equitable breast cancer diagnostic tools, improving patient outcomes and reducing health disparities.

## Dataset

The dataset used for the analysis is The EMory BrEast imaging Dataset (EMBED) (https://arxiv.org/abs/2202.04073) [1]. The dataset comprises of 3.5M screening and diagnostic Mammograms and clinical data about the mammograms. The dataset tries to have a balanced racial demographic sample to help decrease racial bias in AI models.

Our analysis is focussed on the provided clinical data for the mammograms. The complete data has 81776 records and 21 clinical feature columns. The pathology ground truth outcome is labeled as “path severity” which we have used as the target variable for our classification model. Majority of the records have the value for target variable missing, so we have selected 4689 records for which the target variable is present. We selected the following features as our model input patient covariates: “tissueden”, “side”, “numfind”, “total_L_find”, “total_R_find”, “calcnumber”, “ETHNICITY_DESC”, “age_at_study”; and “path_severity as our target variable. The rest of the features had to be dropped due to the majority of the values being null.

The target feature/ outcome variable has 6 different classes. There is an extreme class imbalance present shown below (Table 1). To overcome this, we have used Synthetic Minority Oversampling Technique (SMOTE) oversampling technique to handle the class imbalance. The augmented data has 2132 records for each class. The dataset is split into train (70%) and validation (30%) set. The performance metrics used are: precision, recall and f1-score.

**Table 1.**

| Class Label | Value Count |
| --- | --- |
| 0 | 1069 |
| 1 | 443 |
| 2 | 667 |
| 3 | 39 |
| 4 | 2132 |
| 5 | 76 |

## Model & Results

Instead of doing multi-class classification of the target variable, we have trained 6 binary classification models in order to help with the fairness analysis of the model later. After benchmarking several classification models, we selected Random Forest Classifier. [6] The performance metrics for each model are attached below (Fig.1, Fig.2, Fig.3, Fig.4, Fig.5, Fig.6).

**Fig. 1.**
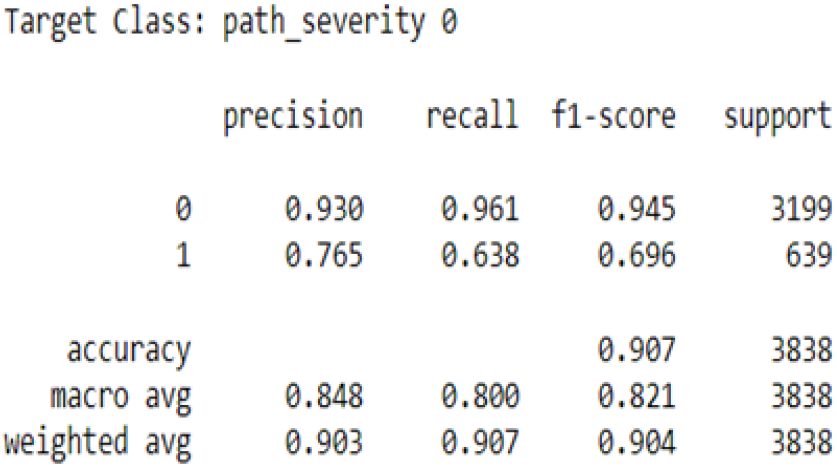

**Fig. 2.**
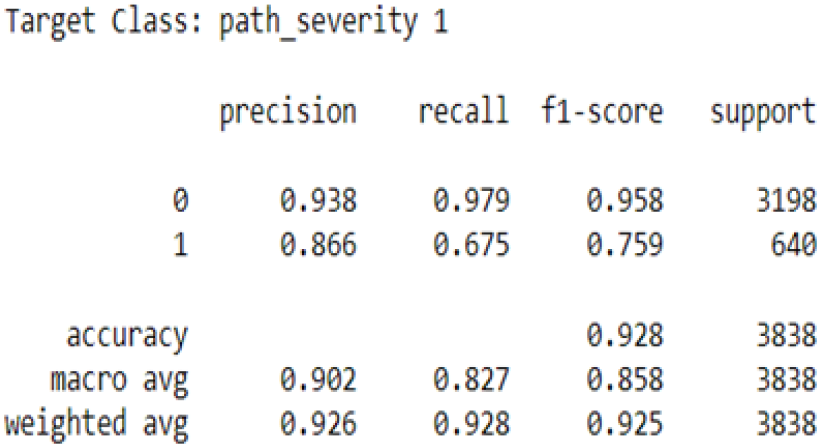

**Fig. 3.**
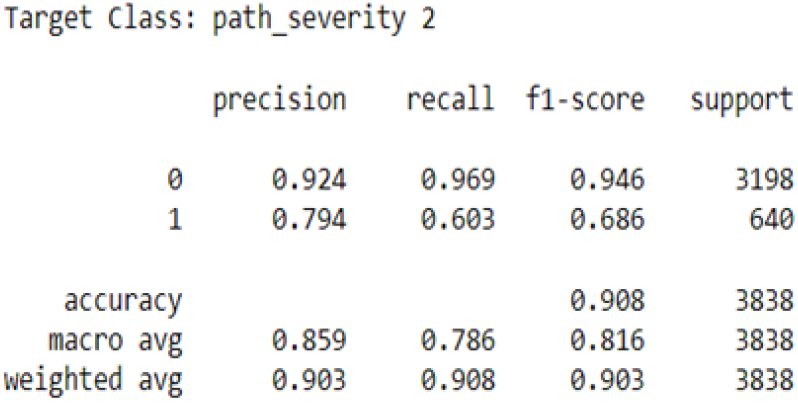

**Fig. 4.**
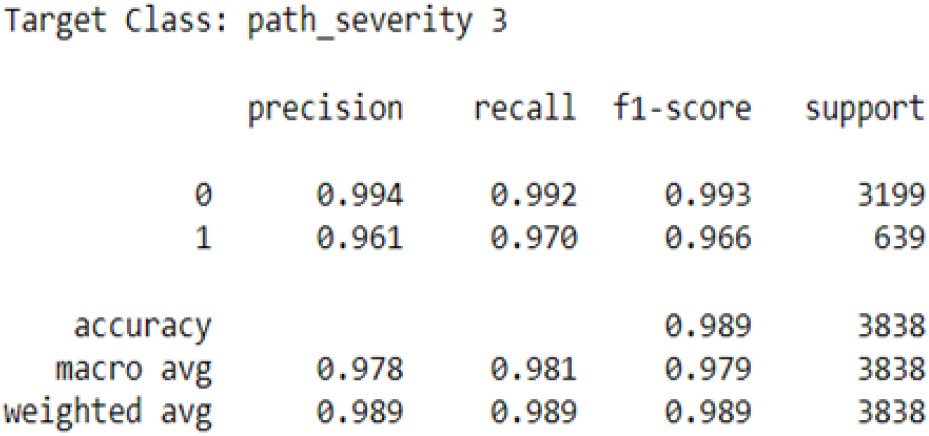

**Fig. 5.**
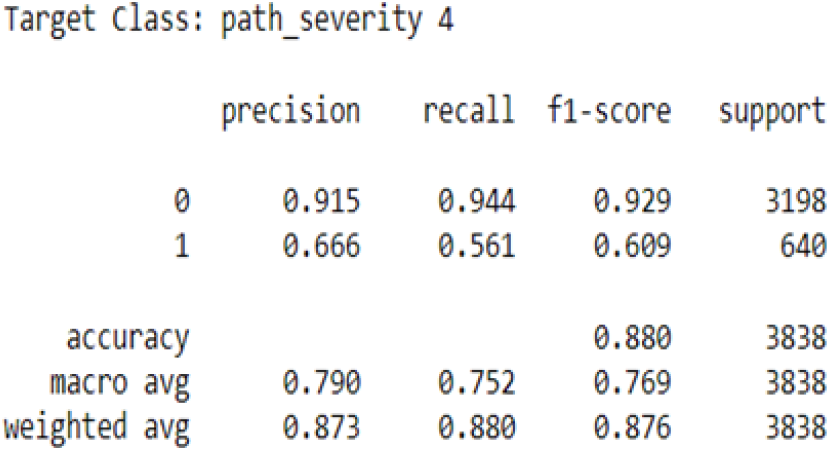

**Fig. 6.**
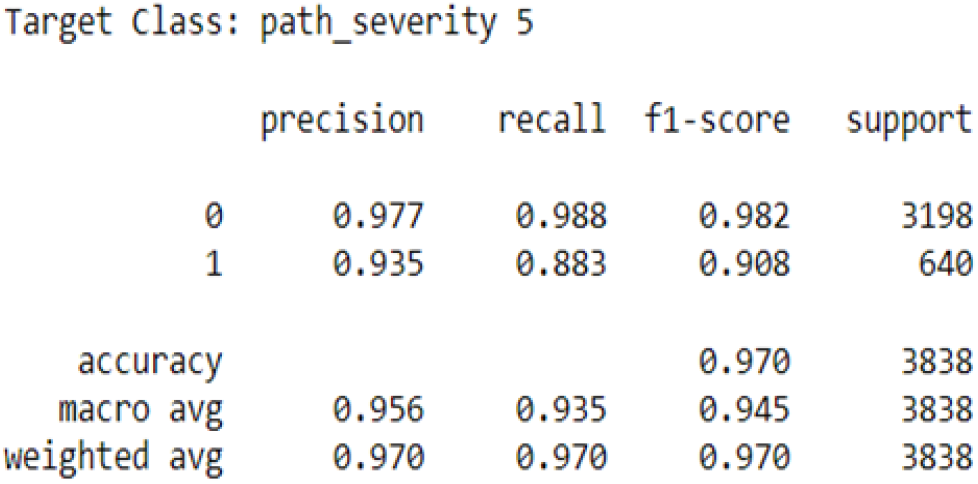

The models performed well with an average accuracy of 93.03 %.

### Fairness Analysis

We have used the following metrics to assess the fairness of the above trained ML models.

Demographic parity: Demographic parity in machine learning refers to the fairness metric that aims to ensure that the model’s outcomes or predictions are similar across different demographic groups in terms of positive outcomes. It requires that the proportion of positive outcomes is similar across different groups, regardless of other factors.

Formula: **(TP + FP)** [4]

Proportional parity: Proportional parity in machine learning is a fairness metric that aims to ensure that the model’s outcomes or predictions are proportional across different demographic groups. It requires that the proportion of positive outcomes is similar across different groups, relative to the proportion of positive outcomes in the population as a whole.

Formula: **(TP + FP) / (TP + FP + TN + FN)** [4]

Equalized odds: Equalized odds in machine learning is a fairness metric that aims to ensure that the model’s outcomes or predictions are accurate and consistent across different demographic groups. It requires that the true positive rate and false positive rate are similar across different groups, ensuring that the model does not unfairly favor or discriminate against any particular group.

Formula: **TP / (TP + FN)** [4]

Predictive rate parity: Predictive rate parity in machine learning is a fairness metric that aims to ensure that the model’s outcomes or predictions are consistent across different demographic groups in terms of predictive accuracy. It requires that the positive predictive value is similar across different groups, ensuring that the model’s accuracy is not biased towards any particular group.

Formula: **TP / (TP + FP)** [4]

Accuracy parity: Accuracy parity in machine learning is a fairness metric that aims to ensure that the model’s outcomes or predictions are consistent across different demographic groups in terms of overall accuracy. It requires that the model’s accuracy is similar across different groups, ensuring that the model is fair and equitable for everyone.

Formula: **(TP + TN) / (TP + FP + TN + FN)** [4]

False negative rate parity: False negative rate parity in machine learning is a fairness metric that aims to ensure that the model’s outcomes or predictions are consistent across different demographic groups in terms of false negative rates. It requires that the model’s false negative rate is similar across different groups, ensuring that the model does not unfairly harm any particular group.

Formula: **FN / (TP + FN)** [4]

False positive rate parity: False positive rate parity in machine learning is a fairness metric that aims to ensure that the model’s outcomes or predictions are consistent across different demographic groups in terms of false positive rates. It requires that the model’s false positive rate is similar across different groups, ensuring that the model does not unfairly favor any particular group.

Formula: **FP / (TN + FP)** [4]

Negative predictive value parity: Negative predictive value parity in machine learning is a fairness metric that aims to ensure that the model’s outcomes or predictions are consistent across different demographic groups in terms of negative predictive values. It requires that the model’s negative predictive value is similar across different groups, ensuring that the model’s accuracy is not biased towards any particular group.

Formula: **TN / (TN + FN)** [4]

Specificity parity: Specificity parity in machine learning is a fairness metric that aims to ensure that the model’s outcomes or predictions are consistent across different demographic groups in terms of specificity values. It requires that the model’s specificity is similar across different groups, ensuring that the model does not unfairly discriminate against any particular group.

Formula: **TN / (TN + FP)** [4]

Matthew’s correlation coefficient comparison: Matthews correlation coefficient comparison in machine learning is a fairness metric that measures the correlation between the actual outcomes and the model’s predictions, taking into account true positive, true negative, false positive, and false negative rates. It is used to compare the performance of different models and ensure fairness across different demographic groups.

Formula: **(TP×TN-FP×FN)/**v**((TP+FP)×(TP+FN)×(TN+FP)×(TN+FN))** [4]

Our dataset contains samples from 3 racial demographics: African Americans, Caucasians, and Asians. To analyze the model’s bias, we segregated the validation set based on racial demographics and used fairness metrics such as Demographic Parity, Proportional parity, etc. The fairness metrics results for each model are attached below (Table 2, Table 3, Table 4).

**Table 2.** African American Demographic

|  | Metric | path_severity_0 | path_severity_1 | path_severity_2 | path_severity_3 | path_severity_4 | path_severity_5 |
| --- | --- | --- | --- | --- | --- | --- | --- |
| 0 | FP | 57.00 | 41.00 | 47.00 | 19.00 | 99.00 | 22.00 |
| 1 | FN | 147.00 | 276.00 | 247.00 | 392.00 | 208.00 | 337.00 |
| 2 | TP | 1874.00 | 1826.00 | 1794.00 | 1797.00 | 1759.00 | 1815.00 |
| 3 | TN | 152.00 | 87.00 | 142.00 | 22.00 | 164.00 | 56.00 |
| 4 | Demographic_parity | 1931.00 | 1867.00 | 1841.00 | 1816.00 | 1858.00 | 1837.00 |
| 5 | Proportional_parity | 0.87 | 0.84 | 0.83 | 0.81 | 0.83 | 0.82 |
| 6 | Predictive_rate_parity | 0.97 | 0.98 | 0.97 | 0.99 | 0.95 | 0.99 |
| 7 | Equalized_odds | 0.93 | 0.87 | 0.88 | 0.82 | 0.89 | 0.84 |
| 8 | Accuracy_parity | 0.91 | 0.86 | 0.87 | 0.82 | 0.86 | 0.84 |
| 9 | False_negative_rate_parity | 0.07 | 0.13 | 0.12 | 0.18 | 0.11 | 0.16 |
| 10 | False_positive_rate_parity | 0.27 | 0.32 | 0.25 | 0.46 | 0.38 | 0.28 |
| 11 | Negative_predictive_value_parity | 0.51 | 0.24 | 0.37 | 0.05 | 0.44 | 0.14 |
| 12 | Specificity_parity | 0.73 | 0.68 | 0.75 | 0.54 | 0.62 | 0.72 |
| 13 | Matthews_correlation_coefficient_comparison | 0.56 | 0.35 | 0.46 | 0.12 | 0.45 | 0.27 |

**Table 3.** Caucasian American Demographic

|  | Metric | path_severity_0 | path_severity_1 | path_severity_2 | path_severity_3 | path_severity_4 | path_severity_5 |
| --- | --- | --- | --- | --- | --- | --- | --- |
| 0 | FP | 60.00 | 27.00 | 45.00 | 12.00 | 67.00 | 15.00 |
| 1 | FN | 212.00 | 140.00 | 124.00 | 173.00 | 131.00 | 201.00 |
| 2 | TP | 1015.00 | 1150.00 | 1124.00 | 1194.00 | 1080.00 | 1141.00 |
| 3 | TN | 99.00 | 69.00 | 93.00 | 7.00 | 108.00 | 29.00 |
| 4 | Demographic_parity | 1075.00 | 1177.00 | 1169.00 | 1206.00 | 1147.00 | 1156.00 |
| 5 | Proportional_parity | 0.78 | 0.85 | 0.84 | 0.87 | 0.83 | 0.83 |
| 6 | Predictive_rate_parity | 0.94 | 0.98 | 0.96 | 0.99 | 0.94 | 0.99 |
| 7 | Equalized_odds | 0.83 | 0.89 | 0.90 | 0.87 | 0.89 | 0.85 |
| 8 | Accuracy_parity | 0.80 | 0.88 | 0.88 | 0.87 | 0.86 | 0.84 |
| 9 | False_negative_rate_parity | 0.17 | 0.11 | 0.10 | 0.13 | 0.11 | 0.15 |
| 10 | False_positive_rate_parity | 0.38 | 0.28 | 0.33 | 0.63 | 0.38 | 0.34 |
| 11 | Negative_predictive_value_parity | 0.32 | 0.33 | 0.43 | 0.04 | 0.45 | 0.13 |
| 12 | Specificity_parity | 0.62 | 0.72 | 0.67 | 0.37 | 0.62 | 0.66 |
| 13 | Matthews_correlation_coefficient_comparison | 0.34 | 0.43 | 0.47 | 0.08 | 0.45 | 0.24 |

**Table 4.**

Asian Demographic
|  | Metric | path_severity_0 | path_severity_1 | path_severity_2 | path_severity_3 | path_severity_4 | path_severity_5 |
| --- | --- | --- | --- | --- | --- | --- | --- |
| 0 | FP | 1.00 | 5.00 | 4.00 | 0.0 | 12.00 | 0.00 |
| 1 | FN | 14.00 | 6.00 | 2.00 | 0.0 | 15.00 | 0.00 |
| 2 | TP | 56.00 | 56.00 | 64.00 | 78.0 | 37.00 | 77.00 |
| 3 | TN | 7.00 | 11.00 | 8.00 | 0.0 | 14.00 | 1.00 |
| 4 | Demographic_parity | 57.00 | 61.00 | 68.00 | 78.0 | 49.00 | 77.00 |
| 5 | Proportional_parity | 0.73 | 0.78 | 0.87 | 1.0 | 0.63 | 0.99 |
| 6 | Predictive_rate_parity | 0.98 | 0.92 | 0.94 | 1.0 | 0.76 | 1.00 |
| 7 | Equalized_odds | 0.80 | 0.90 | 0.97 | 1.0 | 0.71 | 1.00 |
| 8 | Accuracy_parity | 0.81 | 0.86 | 0.92 | 1.0 | 0.65 | 1.00 |
| 9 | False_negative_rate_parity | 0.20 | 0.10 | 0.03 | 0.0 | 0.29 | 0.00 |
| 10 | False_positive_rate_parity | 0.12 | 0.31 | 0.33 | NaN | 0.46 | 0.00 |
| 11 | Negative_predictive_value_parity | 0.33 | 0.65 | 0.80 | NaN | 0.48 | 1.00 |
| 12 | Specificity_parity | 0.88 | 0.69 | 0.67 | NaN | 0.54 | 1.00 |
| 13 | Matthews_correlation_coefficient_comparison | 0.46 | 0.58 | 0.69 | NaN | 0.24 | 1.00 |

To optimize our model to play fair for all racial demographics. We take help from an open-source project called fairlearn (https://github.com/fairlearn/fairlearn) [2]. We use the Threshold Optimizer algorithm from the package to equalize the odds between different groups of users of our model. The Threshold Optimizer is based on a paper called “Equality of Opportunity in Supervised Learning” (https://arxiv.org/abs/1610.02413) [3]. To compare the results of the original and fair model, we calculate the fairness metrics of both models, find out the confidence intervals and use histograms to visualize any disparity.

To obtain confidence intervals, we first resample 70% of the data from the original dataset to be used as the training set and the remaining records to be used as the validation set. Then both the model and fair model are trained on the training dataset and fairness metrics are calculated. In total 12 models are trained 6 normal and 6 fair models, each model a binary classification for one of the six target variables. The above steps are iterated 1000 times. Then the collected data is used to plot histograms and calculate the confidence interval for each metric for each model.

Confidence interval plots were generated for the above models. Below attached are the figures which show low overlap between the fair and regular models, which signifies there is a statistical difference between the performance of models. The plots show that the fair model might be less accurate but will not punish any particular demographic.

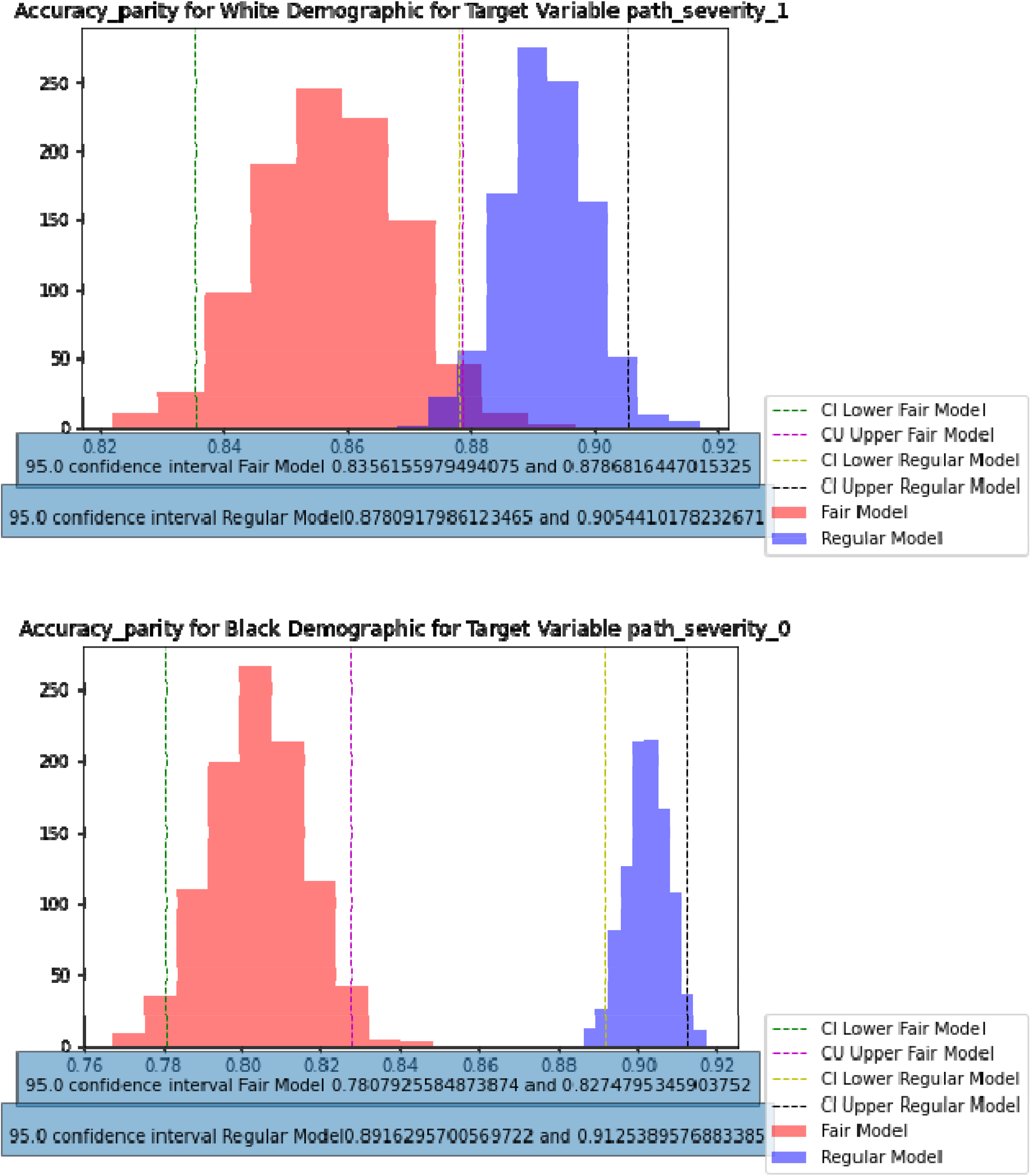

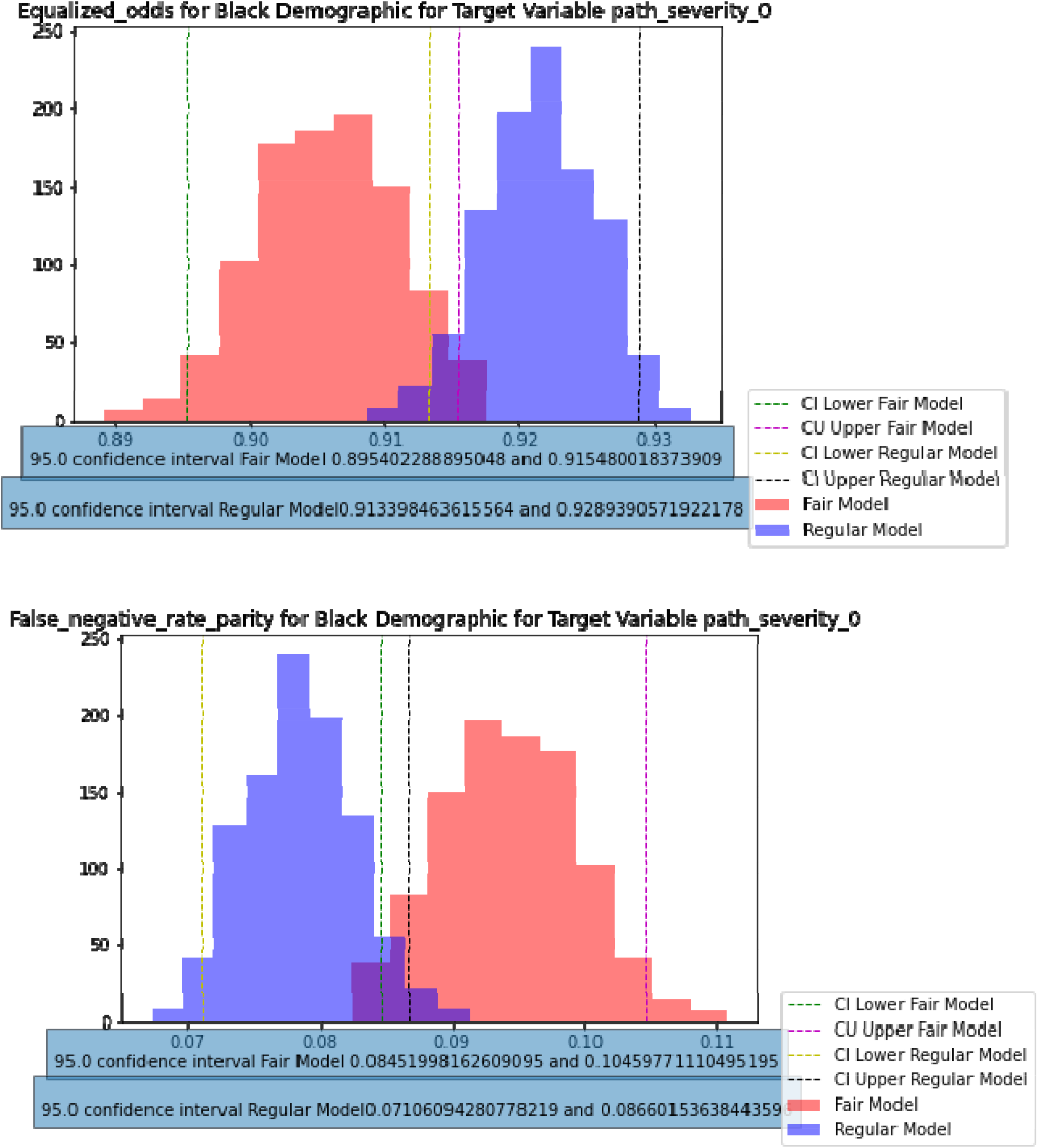

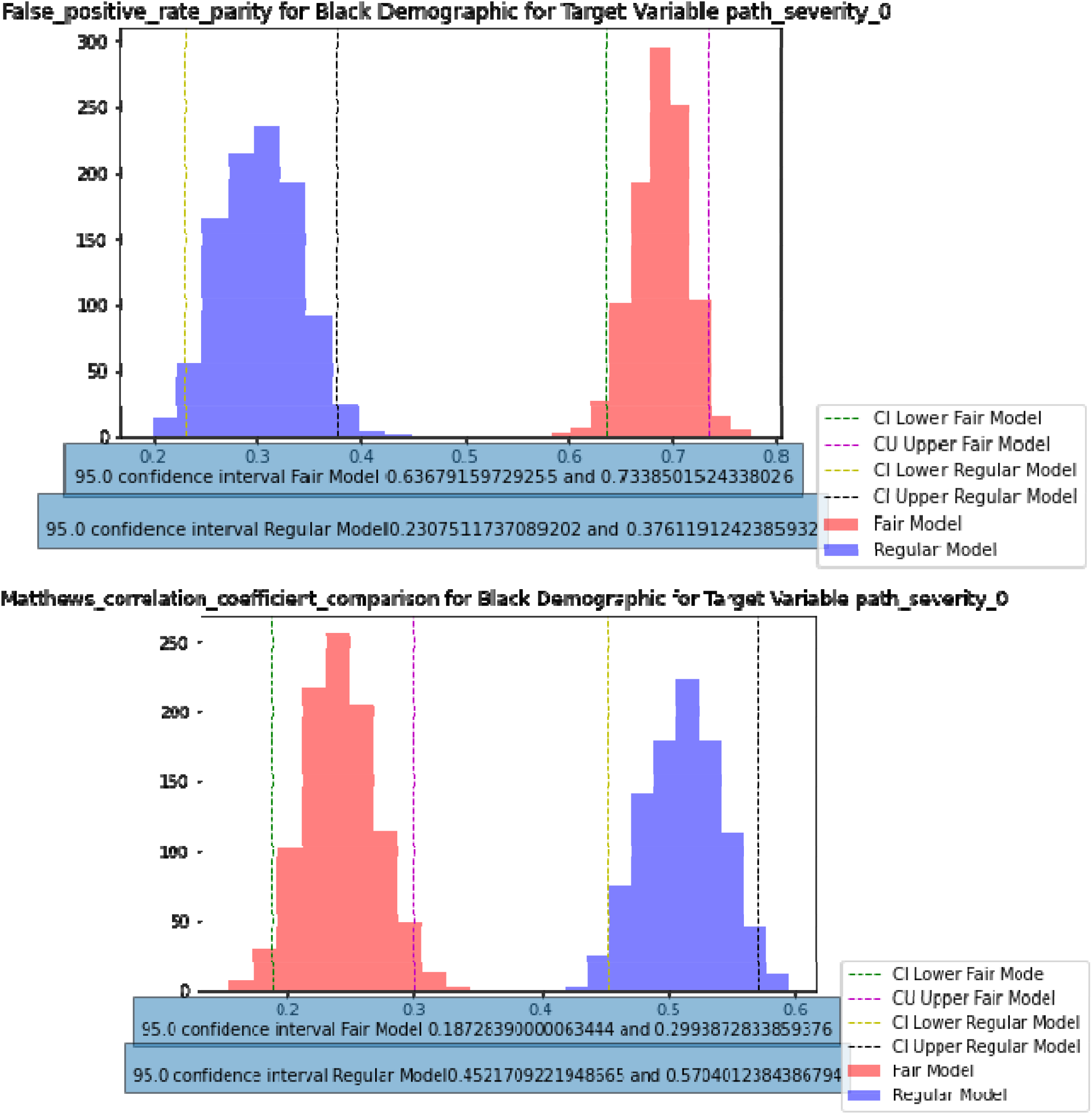

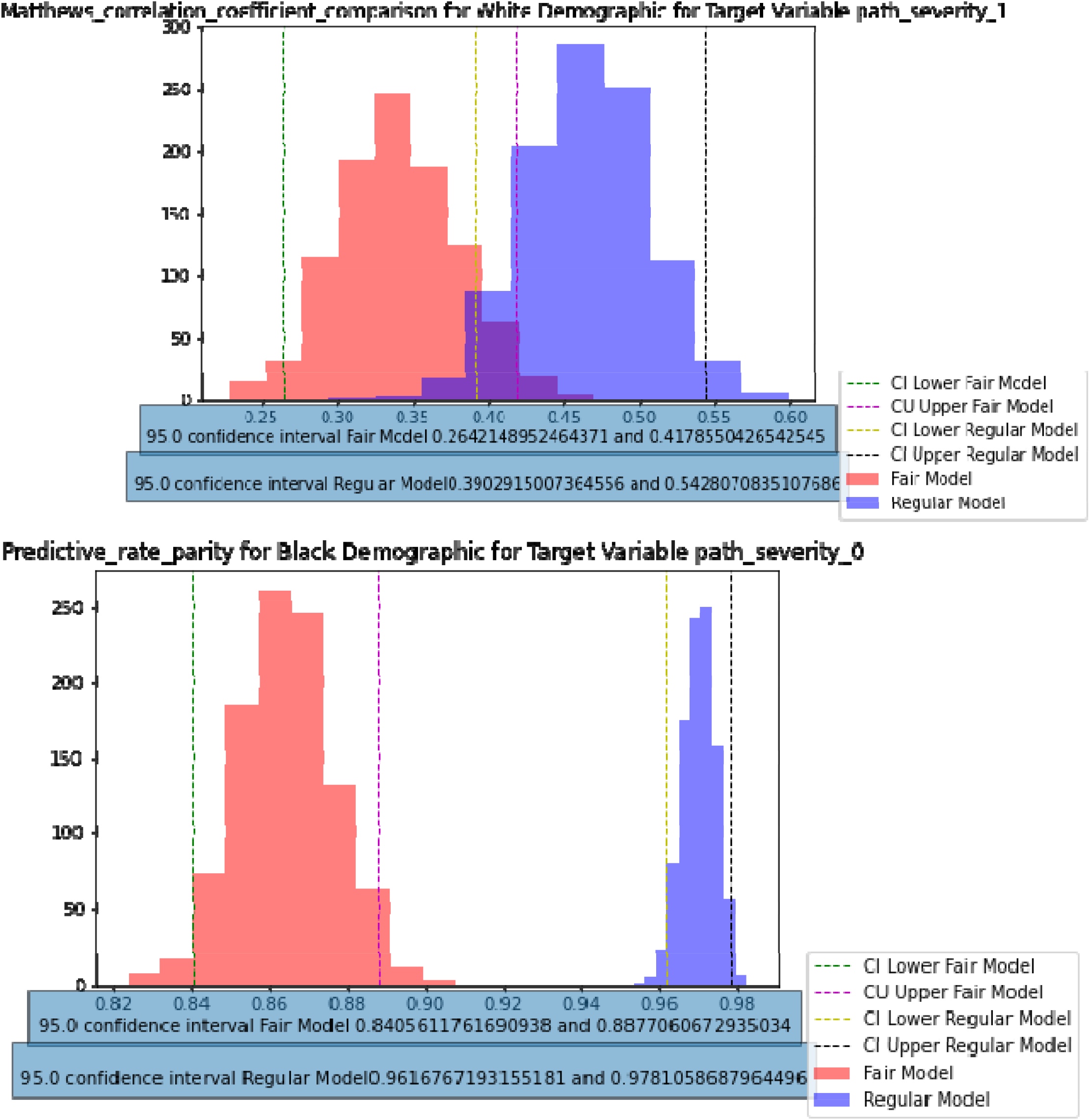

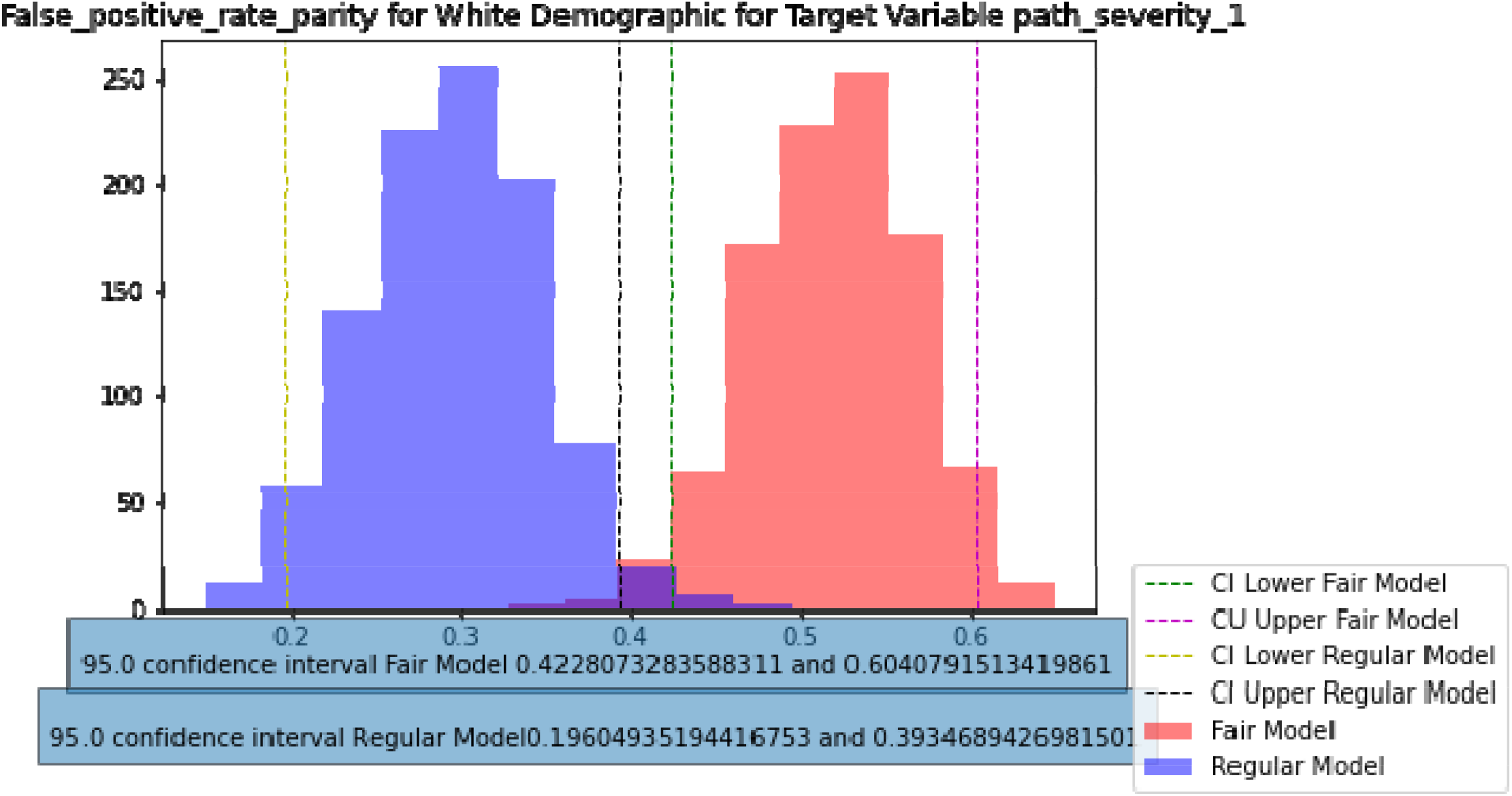

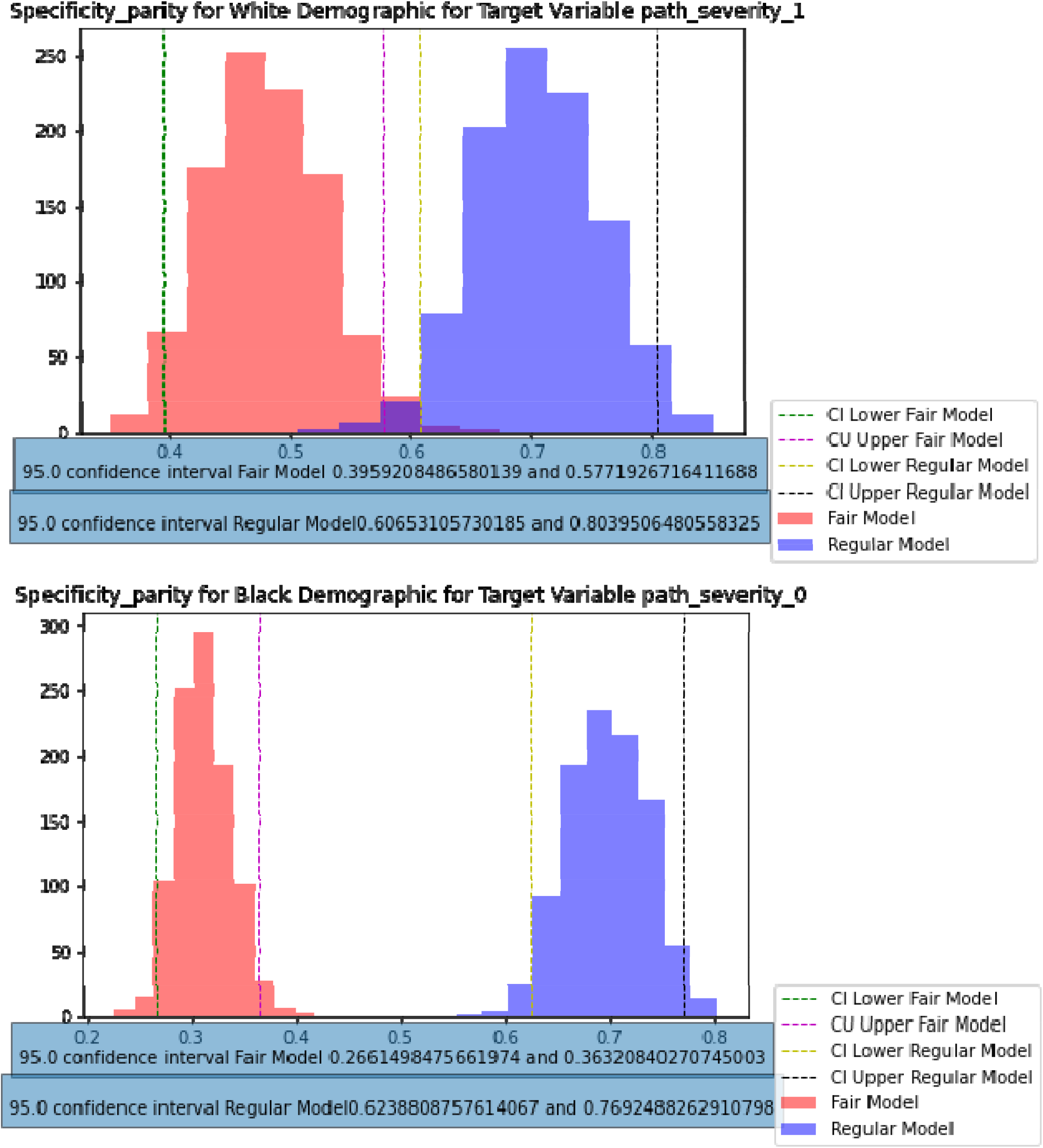

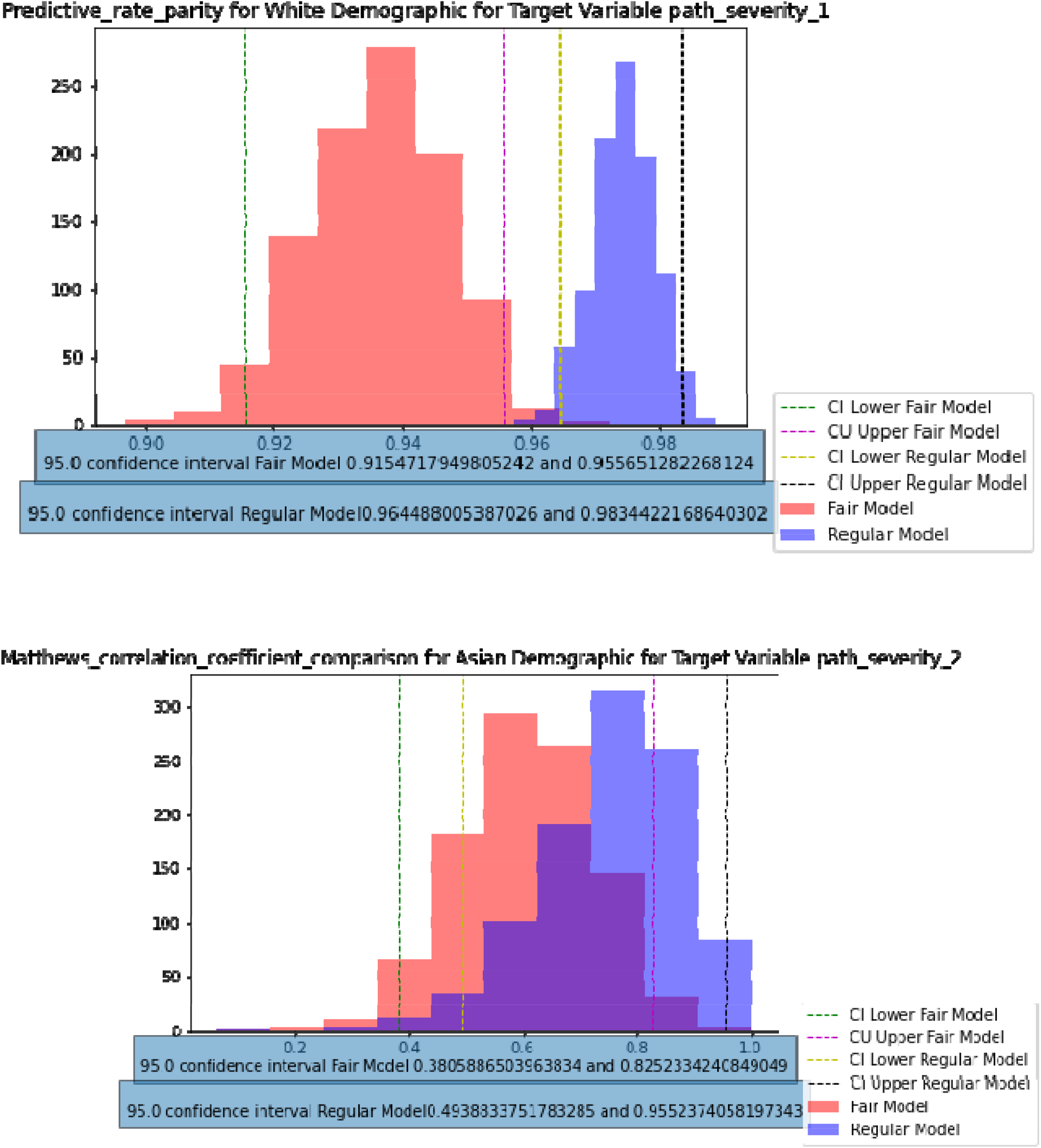

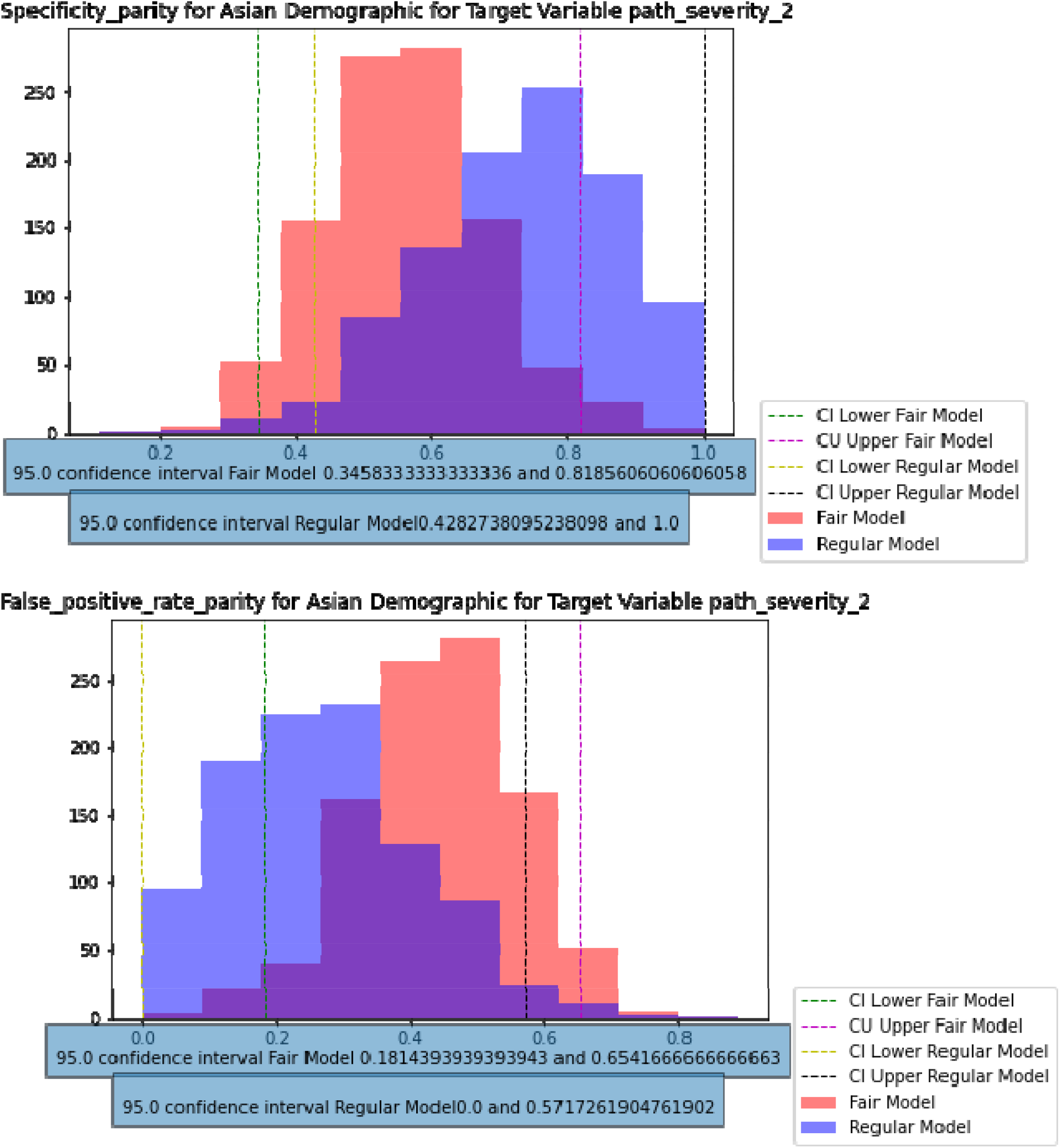

| Target Variable | Random Forest Accuracy | Fair Model Accuracy |
| --- | --- | --- |
| path_severity_0 | 0.9017 | 0.8410 |
| path_severity_1 | 0.9296 | 0.9161 |
| path_severity_2 | 0.9090 | 0.9015 |
| path_severity_3 | 0.9887 | 0.9627 |

## Data Availability

All data is available from the EMBED website upon request for non commercial/academic use

## Appendix 1: Multimodal Image Classification

### Introduction

In addition to our machine learning-based predictive modeling on clinical data from the Emory Breast Imaging Dataset (EMBED) [1] and model fairness analysis, we also propose a multi-modal classification approach that combines both images and clinical features to predict the target variable of path severity.

Breast cancer diagnosis and prognosis typically rely on mammography images and clinical data, such as patient age, family history, and biopsy results. However, these approaches are limited by their reliance on either images or clinical data alone. By combining both modalities, we can potentially improve the accuracy and reliability of breast cancer diagnosis and prognosis.

By integrating both modalities, we hope to develop a more comprehensive and accurate breast cancer diagnostic and prognostic tool that can ultimately improve patient outcomes. This multi-modal approach can potentially reduce the need for invasive procedures and provide more personalized treatment recommendations for patients with breast cancer.

#### Dataset

The ‘path_severity’ target variable is only present for 4788 rows and there is a huge imbalance as mentioned above already. So we decided to only do binary classification for target labels 0 and 4.The value count of both classes is mentioned below

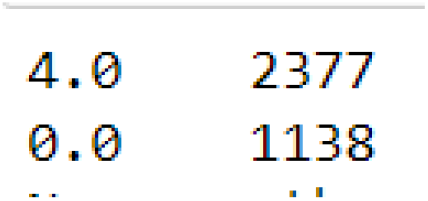

We selected the following features as our input for the ANN model (clinical features): “tissueden”, “side”, “numfind”, “total_L_find”, “total_R_find”, “calcnumber”, “ETHNICITY_DESC”, “age_at_study” and “path_severity: as our target variable. The rest of the features had to be dropped due to the majority of the values being null. The corresponding mammogram images for each record were used as input for our CNN model.

### Model & Results

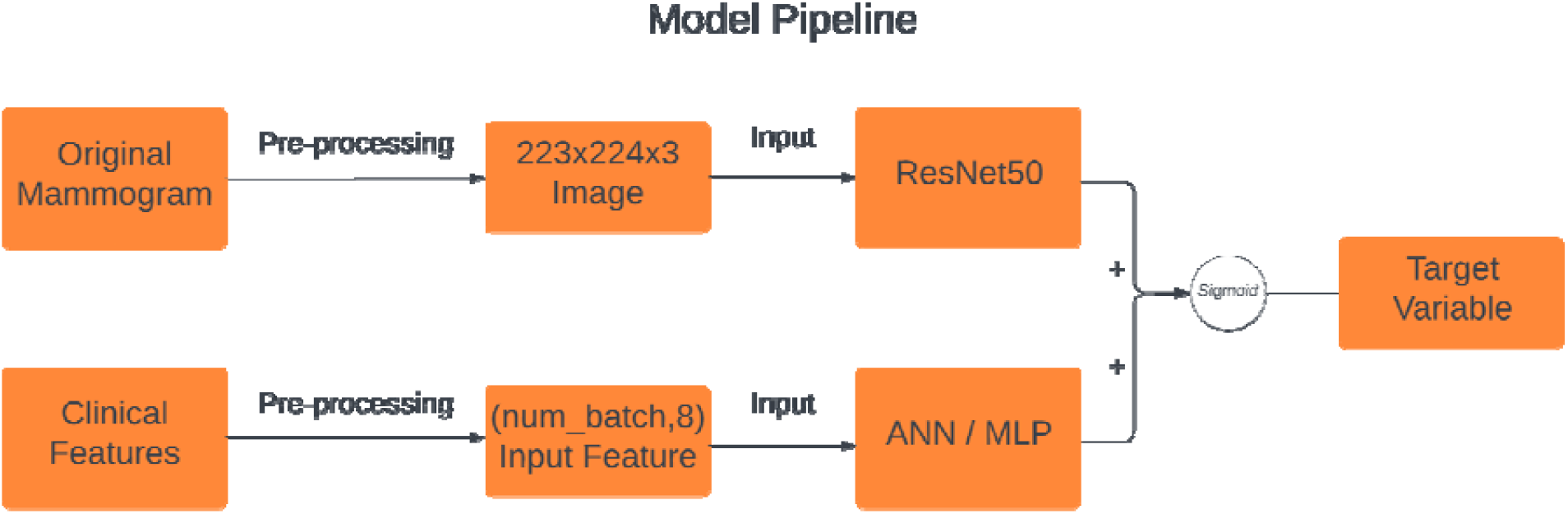

|  | Loss | Accuracy |
| --- | --- | --- |
| Train | 0.588 | 68.85 |
| Validation | 0.586 | 68.22 |

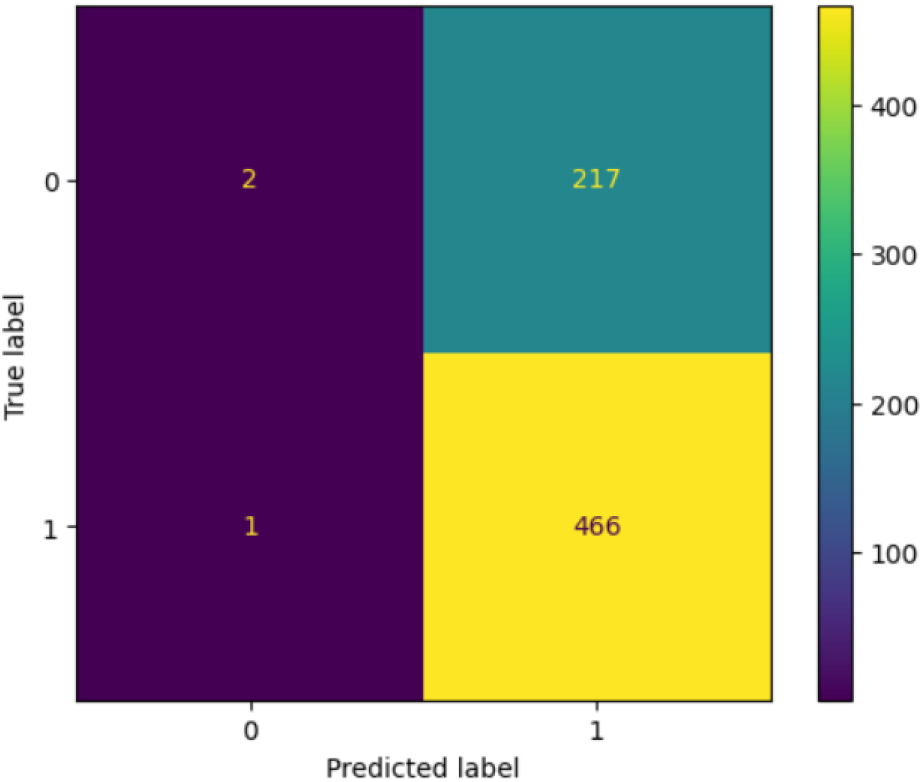

#### Dataset

The ‘path_severity’ target variable is only present for 4788 rows and there is a huge imbalance as mentioned above already. So we decided to only do binary classification for target labels 0 and 4. The value count of both classes is mentioned below

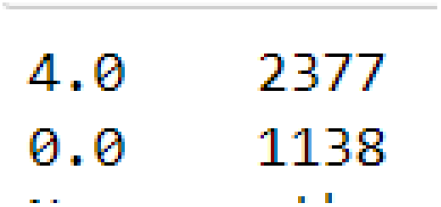

This time we only used mammogram images to perform binary classification

### Model & Results

We trained the model for 20 epochs.

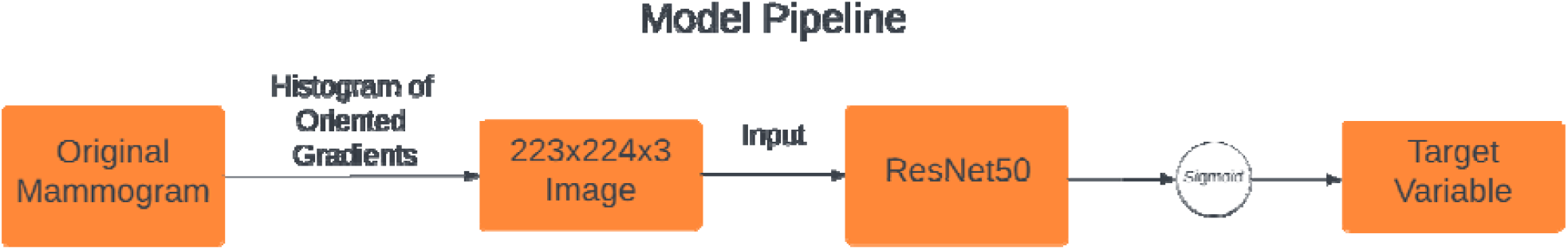

|  | Loss | Accuracy |
| --- | --- | --- |
| Train | 0.481 | 67.60 |
| Validation | 0.674 | 67.520 |

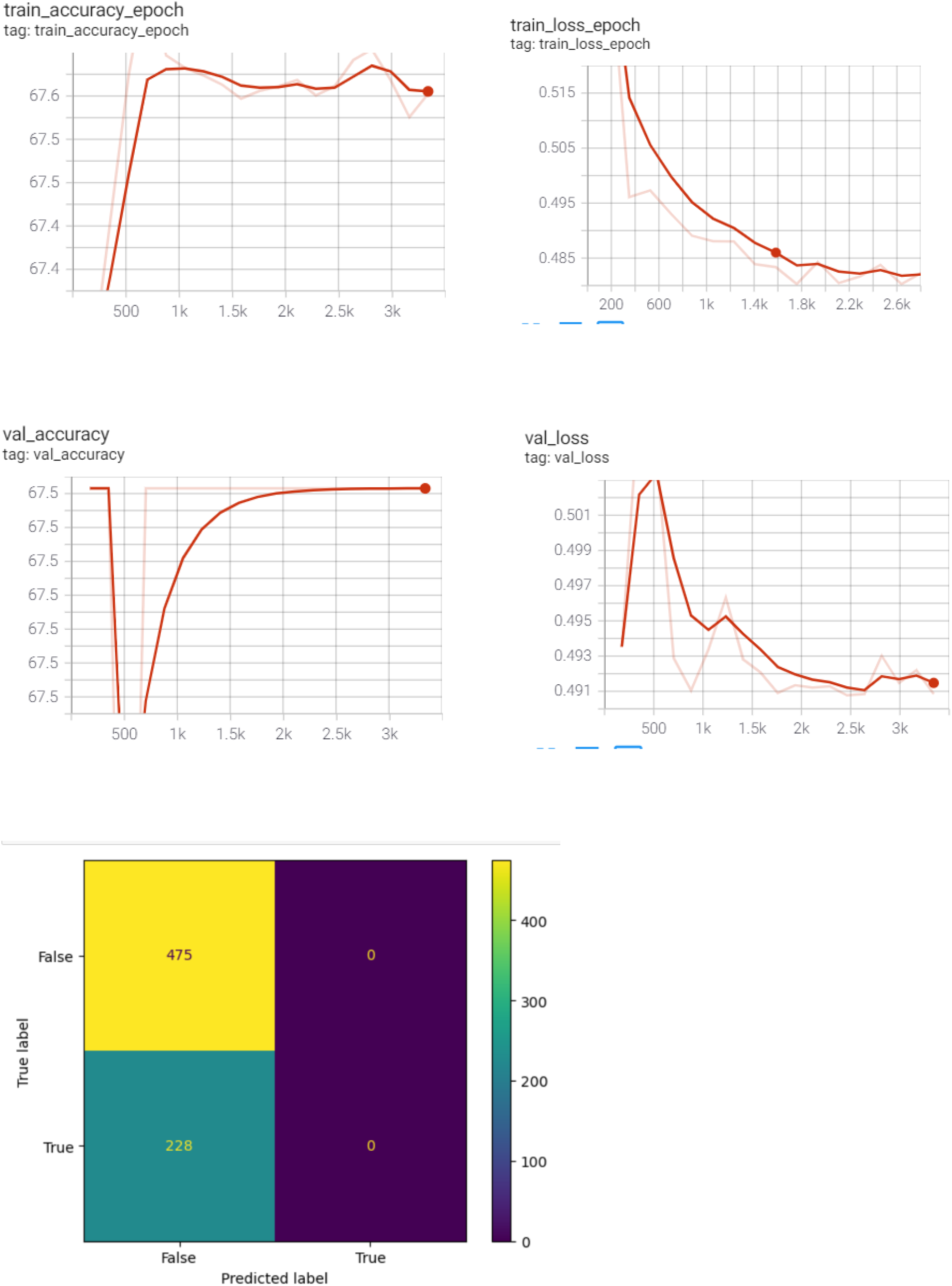

Despite our best efforts, we encountered challenges in implementing our multi-modal classification approach for predicting path severity in breast cancer using both images and clinical features from the Emory Breast Imaging Dataset (EMBED) [1]. We encountered difficulties in training the model, with the loss function not reducing and poor accuracy results. Our attempts to use a combination of convolutional neural networks (CNNs) and Artificial Neural Networks (ANN / MLP) did not yield the expected results. While multi-modal approaches have shown promise in improving the performance of the model, the complexity and heterogeneity of the data in EMBED may have presented additional challenges in implementing a successful multi-modal approach.

Despite these challenges, our work highlights the importance of continued research and development in multi-modal approaches to breast cancer diagnosis and prognosis. While our approach may not have yielded the desired results, our work provides valuable insights into the challenges and opportunities presented by multi-modal data analysis in the field of breast cancer research. In the future, we believe that further research and development in multi-modal approaches can lead to more accurate and reliable breast cancer diagnostic and prognostic tools that can ultimately improve patient outcomes. While our work may not have achieved the desired results, we remain optimistic about the potential of multi-modal approaches in the fight against breast cancer.

